# Associations Between Cannabis Use and Sleep Outcomes in a Sample of Pregnant Women

**DOI:** 10.64898/2026.10.30.26366959

**Authors:** Aaron J. Gorelik, Ethan Kinstler, Alicia Cuñado, Rebecca C. Cox, Anna Constantino-Pettit, Shelby D. Leverett, Alexander S. Hatoum, Shannon N. Lenze, Christopher D. Smyser, Cynthia E. Rogers, Arpana Agrawal, Ryan Bogdan, Emma C. Johnson

**Affiliations:** Department of Psychological and Brain Sciences, Washington University in St Louis, St Louis, Missouri; Department of Psychiatry, Washington University in St Louis, School of Medicine, St Louis, Missouri; Department of Neurology, Washington University in St Louis, St Louis, Missouri

**Keywords:** Cannabis use, pregnancy, sleep

## Abstract

**Importance:** Both sleep difficulties and cannabis use during pregnancy (CUP) are common and associated with adverse birth and offspring outcomes. Sleep problems are a commonly reported motive for cannabis use during pregnancy.

**Objective:** Estimate associations between cannabis use during pregnancy and common sleep difficulties across trimesters.

**Design, Setting, and Participants:** This cohort study recruited pregnant women with lifetime cannabis use who used (n=298) and did not use (n=143) cannabis during pregnancy. Women who reported heavy episodic alcohol consumption or use of other illicit substances were excluded. Some women tested positive for cotinine in urine drug screens.

**Exposures:** Cannabis use was assessed at each trimester using self-report and urine drug screening. Presence of the nicotine metabolite, cotinine, was measured via urine drug screening at each trimester. Cannabis use during pregnancy (CUP) status was defined separately for each trimester as use versus no use and was treated as a time-varying exposure. The nicotine metabolite cotinine was included as a covariate in all models to account for nicotine exposure. Motivation for cannabis use during pregnancy was assessed during the first trimester.

**Main Outcomes and Measures:** Insomnia symptoms (Women’s Health Initiative Insomnia Rating Scale), daytime sleepiness (Epworth Sleepiness Scale), subjective sleep quality (Pittsburgh Sleep Quality Index single item), and restless legs symptoms were assessed at each trimester.

**Results:** A *CUP Status* x *Trimester* interaction was observed for insomnia symptoms (F(2,737.81)=4.98, *P*_FDR_=0.03) wherein use during the third trimester was associated with heightened insomnia. Women reporting sleep difficulties as a motive for use reported worse insomnia symptoms (F(1, 223)=13.09, *P*_FDR_=0.001) and worse sleep quality (F(1, 219)=7.86, *P*_FDR_=0.01), relative to those using for other reasons.

**Conclusion:** Elevated third trimester insomnia symptoms among pregnant women using cannabis compared to women who did not use cannabis suggest a potential trimester-sensitive association. Despite perceptions that cannabis improves sleep, women who reported sleep as their motive for CUP during the first trimester experienced greater insomnia symptoms and poorer sleep quality across pregnancy than those using cannabis for other reasons. Additional studies are needed to determine whether sleep-related difficulties precede cannabis use, follow it, are modified by it, or represent an epiphenomenon.

**KEY POINTS:** *Question:* Is cannabis use during pregnancy associated with maternal sleep problems?

*Findings:* Within this study of 441 pregnant women, cannabis use was associated with higher third trimester insomnia symptoms but not with other sleep outcomes. Women using cannabis to aid with sleep difficulties had worse insomnia symptoms and poorer sleep quality than those using for other reasons.

*Meaning:* Cannabis use in late pregnancy may mark periods of greater insomnia symptom burden, suggesting that sleep problems during pregnancy may be an important clinical context for understanding cannabis use during pregnancy.

## INTRODUCTION

Despite its association with adverse offspring outcomes,^1,2^ cannabis use during pregnancy (CUP) has become increasingly common, with rates increasing more than 2-fold between 2002-2020.^3^ Sleep is commonly disrupted during pregnancy,^4,5^ and addressing sleep problems and other factors that may influence sleep (e.g., nausea, pain, stress, depression) are common motives for cannabis use across multiple populations,^6^ including pregnant women.^7,8^ CUP to address sleep challenges may be motivated by the notion that cannabis is a safer alternative to prescription-based sleep medications.^9^

Despite widespread perceptions that cannabis improves sleep,^10^ there is equivocal evidence that cannabis is associated with both better and worse sleep outcomes^10–14^ and a meta-analysis of medical cannabis treatment of insomnia found low certainty evidence of little to no impact on sleep.^15^ While many studies examine the relationship between lifetime or current cannabis use and aspects of sleep,^16–19^ investigations of use during pregnancy are limited; indeed, we are only aware of two studies which measured use and sleep during pregnancy. One found no association between use and sleep quality^20^ while the other found greater insomnia severity, but no differences in sleep quality, specifically among those using cannabis for sleep versus other reasons.^21^

Here, in a sample of 441 pregnant women, we estimated associations between CUP and the following sleep problems: insomnia symptoms, daytime sleepiness, sleep quality, and restless legs symptoms. We examined whether associations between CUP and these sleep outcomes differed across trimesters, and whether frequency of use and motive for use were associated with sleep outcomes. We hypothesized that CUP would be associated with worse sleep overall across all assessed outcomes and that sleep would worsen across trimesters.

## METHODS

### Participants

The ongoing Cannabis Use During Early Life and Development (CUDDEL) study is a longitudinal cohort study of prenatal cannabis exposure and offspring brain and behavior development.^8,22^ Pregnant women (n=441) who did (n=298) and did not use cannabis (n=143) during pregnancy were recruited from the Barnes-Jewish Hospital Pregnancy Clinic in Saint Louis, MO from 07/2019-01/2024. See **eMethods** for complete inclusion criteria. At each trimester clinical care visit, participants completed questionnaires and provided urine for research drug testing. All participants provided written informed consent to a protocol approved by the Washington University institutional review board **(eMethods)**. There were no institutional or state reporting requirements for cannabis use during pregnancy reported in a research setting. The final maximum analytic sample consisted of 411 pregnant women, 278 of whom (67.6%) reported or tested positive for cannabis during pregnancy (see below). This study followed the Strengthening the Reporting of Observational Studies in Epidemiology ([STROBE](http://www.equator-network.org/reporting-guidelines/strobe/))^23^ reporting guidelines.

### Measures

#### Cannabis Use During Pregnancy

Trimester-specific cannabis use during pregnancy (CUP) status, which could vary within individuals across trimesters, was coded as positive for participants self-reporting cannabis use *<u>or</u>* having urine positivity during study visits (n_T1_=261 [60.6%], n_T2_=212 [51.1%], n_T3_=178 [44.7%]) and negative when participants reported no use *<u>and</u>* tested negative in available urine samples at a given trimester visit (n_T1_=170 [39.4%], n_T2_=203 [48.9%], n_T3_=220 [55.3%]). Binary CUP Status (cannabis use versus no use) had moderate stability across trimesters: T1-T2 r=.601, T1-T3 r=.521, T2-T3 r=.696. ***Self-report***. Participants reported their frequency of use (e.g., never, 2-4x/month; **eMethods**) at each trimester based on item 1 of the Cannabis Use Disorders Identification Test-Revised^24^ with modifications to assess use during pregnancy, during the first trimester prior to and subsequent to knowledge of pregnancy, and since the prior study visit in trimesters 2 and 3. For analyses, women who self reported no use but who tested positive for at least one trimester were excluded (n=13). Frequency was recoded at each trimester as never (n_T1_=152, n_T2_=168, n_T3_=191), monthly (n_T1_=68, n_T2_=45, n_T3_=41), or at least 2 or more times per month (n_T1_=118, n_T2_=103, n_T3_=82; **eMethods**). ***Urine Drug Testing***. At each clinical-care trimester visit, urine was tested for the cannabis metabolite THC-COOH using commercially available urine drug testing kits (**eMethods**). Concordance between self-reported use and THC positive urine screens was moderate (r=0.7), indicating reasonable agreement.^22^

#### Other Substance Use During Pregnancy

Other substance use during pregnancy was exclusionary. ***Self-report***. At each study visit, participants completed the 3-item alcohol consumption scale from the Alcohol Use Disorders Identification Test^25^ and were queried on their use of tobacco-containing products, other illicit drugs, and off-label use of prescriptions (full list in **eMethods). *Urine Drug Testing***. At each trimester visit, urine was tested for the presence of other substance metabolites (e.g., amphetamines; see full list of substances in **eMethods**), using commercially available drug testing cups (**emethods**). Participants were excluded from the study if they reported use of these substances or tested positive for any of these substances at any trimester assessment. The nicotine metabolite cotinine was tested for in urine at each trimester visit using a commercially available cotinine strip test (**eMethods**). Women who used cannabis and reported no nicotine use, but tested positive for cotinine, were retained within the study as this may be attributable to blunts as a mode of use in this sample and/or second-hand exposure. Among participants who reported cannabis use or tested positive for cannabis during pregnancy, 158 (37.2%) tested positive for cotinine and 267 (62.8%) tested negative. Ever testing positive at any trimester was used as a covariate in analyses. Women without evidence of cannabis use at any trimester visit who tested positive for cotinine were excluded from the study.

#### Sleep

Insomnia symptoms (Women’s Health Initiative Insomnia Rating Scale [WHIIRS],^26^ daytime sleepiness (Epworth Sleepiness Scale [ESS]),^27^ restless legs symptom severity (National Institutes of Health panel consensus criteria),^28^ and sleep quality (Pittsburgh Sleep Quality Index [PSQI] single item)^29^ were assessed at each trimester.

#### Sleep Motive for Cannabis Use During Pregnancy

During T1, individuals reporting CUP were asked about their motives for use during pregnancy with the following response options (multiple responses permitted) based on prior questionnaires including the *Pregnancy Risk Assessment Monitoring Survey*^30^ (e.g., to deal with nausea and vomiting, to help you sleep, etc; see **eMethods** for complete list of options). We categorized participants into those reporting sleep as a motive (n=96) and those not reporting sleep as a motive (n=155).

#### Race & Ethnicity

Participants were asked to self-identify their race during T1 to characterize the racial and ethnic make up of the sample (**eMethods**).

#### Income-to-needs Ratios

Income-to-needs ratios were derived by dividing participant reported total-household-income (assessed during the first trimester) by the federal guideline for poverty (from the same year the participant reported) given reported family size. Lower income-to-needs ratios denote greater socioeconomic disadvantage. In analytic sample, income-to-needs ratios averaged 1.21±0.99 (Range: 0.34-9.72).

#### Statistical Analysis

Linear mixed-effects regressions were used to estimate associations of *CUP Status, Trimester* (T1,T2,T3), and their interaction (i.e., *CUP Status* x *Trimester*) for each sleep phenotype, using the ‘lme4’ R (v4.3.2) package (v1.1-30).^31^ We chose to model trimester as a categorical variable and did not analyze longitudinal trajectories because of the variability in timing of assessments within trimester (e.g., one participant may have been assessed closer to the beginning of T1, while another participant may have been assessed in the middle of T1). As such, *CUP Status* was treated as a time-varying, trimester-level exposure, allowing individuals to change *CUP Status* across trimesters. Omnibus effects were evaluated using type II sums of squares due to unbalanced groups.^32^ To account for testing across the four sleep phenotypes, false discovery rate correction was adjusted separately for each model term (i.e., *CUP Status, Trimester, CUP Status* x *Trimester*). Pairwise contrasts were used to characterize significant main and interactive effects using the emmeans R package.^33^ ***Covariates***. Models included urine cotinine positivity at any trimester assessment, mean centered age, and mean centered income-to-needs ratio as fixed effect covariates as well as their interactions with *Trimester* and with *CUP Status* to fully account for potential covariate confounding (i.e., predictor x covariate interactions).^34^ As mental health history and prescription of psychotropic medications obtained from electronic health records were not associated with CUP status, they were not considered as covariates.^8^ Sensitivity analyses evaluated the impact of missing third trimester data (i.e., models rerun with missing third trimester *CUP Status* and sleep data adjusted for as covariates; models rerun excluding participants with missing third trimester data [n_excluded_=50]). Secondary analyses of *Cannabis Use Frequency* (i.e., never, monthly, or at least 2 or more times per month) and *Sleep Motive* (sleep motive reported n=96; not reported n=155) were conducted using separate linear mixed-effects models estimating their main effects, main effects of trimester, and their interaction. An identical FDR correction approach was applied independently to the cannabis use frequency and sleep motive analyses.

## RESULTS

### Sample Characteristics

The sample was majority Black (75.6%) and non-Hispanic (95.6%), with relatively low income-to-needs ratios (indicative of greater resource scarcity; mean=1.22, SD=0.99) and an average age of 26.6 years at their first trimester (**Table 1**). Of the full analytic sample, 286 (67.9%) women reported cannabis use (n=243) and/or tested positive in urine drug screens (n=233) at any trimester with 135 (32.1%) reporting no use and testing negative at every trimester. Among women using cannabis during pregnancy in the analytic sample, 133 (46.8%) did not test positive for cotinine at any trimester assessment, while 151 (53.2%) did.

**Table 1.** Descriptive statistics of the sample across trimesters. *nocan*=negative cannabis use during pregnancy status; *can*=positive cannabis use during pregnancy status. Income-to-needs ratio, race, and ethnicity did not change across trimesters and thus are only reported once. Cotinine positivity was assessed here via urine drug screen only. *Note*. For all statistical analyses CUP Status (i.e., nocan and can) were categorized for each respective trimester.

| **Characteristic** | **Overall**<br>N = 431<sup>1</sup> | **nocan**<br>N = 170<sup>1</sup> | **can**<br>N = 261<sup>1</sup> | **p-value** |
|---|---|---|---|---|
| Race |  |  |  | 0.9<sup>2</sup> |
| Black | 326 | 128 | 198 |  |
| Other | 20 | 7 | 13 |  |
| White | 85 | 35 | 50 |  |
| Ethnicity |  |  |  | >0.9<sup>2</sup> |
| Hispanic | 19 | 8 | 11 |  |
| Non-Hispanic | 412 | 162 | 250 |  |
| Maternal Age | 26.63 (4.96) | 27.25 (5.37) | 26.23 (4.64) | 0.042<sup>3</sup> |
| Income-to-Needs | 1.21 (0.99) | 1.47 (1.41) | 1.04 (0.51) | <0.001<sup>3</sup> |
| Insomnia symptoms (T1) | 8.42 (5.31) | 8.90 (5.29) | 8.10 (5.31) | 0.14<sup>3</sup> |
| Insomnia symptoms (T2) | 7.82 (5.39) | 8.36 (5.35) | 7.46 (5.41) | 0.2<sup>3</sup> |
| Insomnia symptoms (T3) | 8.05 (5.40) | 8.21 (5.13) | 7.95 (5.57) | 0.7<sup>3</sup> |
| Daytime Sleepiness (T1) | 7.85 (5.02) | 8.20 (5.48) | 7.61 (4.68) | 0.3<sup>3</sup> |
| Daytime Sleepiness (T2) | 7.74 (5.18) | 7.52 (4.97) | 7.88 (5.34) | 0.6<sup>3</sup> |
| Daytime Sleepiness (T3) | 7.11 (5.00) | 6.96 (5.11) | 7.19 (4.97) | 0.8<sup>3</sup> |
| Restless Leg (T1) | 0.53 (0.94) | 0.51 (0.89) | 0.54 (0.98) | 0.7<sup>3</sup> |
| Restless Leg (T2) | 0.59 (1.04) | 0.62 (1.04) | 0.57 (1.04) | 0.7<sup>3</sup> |
| Restless Leg (T3) | 0.68 (1.19) | 0.62 (1.14) | 0.70 (1.22) | 0.6<sup>3</sup> |
| Cotinine Positivity (T1) | 104 | 0 | 104 |  |
| Cotinine Positivity (T2) | 90 |  | 90 |  |
| Cotinine Positivity (T3) | 78 |  | 78 |  |

<sup>1</sup>n; Mean (SD)
<sup>2</sup>Pearson's Chi-squared test
<sup>3</sup>Welch Two Sample t-test

### Cannabis Use Across Pregnancy

Trimester-specific cannabis use during pregnancy (CUP) status (i.e., positive self-report or positive urine toxicology) was observed in 249 of 409 women at T1 (60.9%), 197 of 386 women at T2 (51%), and 166 of 375 at T3 (44.3%). Cannabis use differed significantly across trimesters; Cochran’s Q(2)=46, *P*<0.001. Holm-adjusted pairwise McNemar tests indicated significant trimester-level decline in women who used cannabis during pregnancy from T1 to T2, χ^2^(1) = 17.81, *P*_adjusted_< .001; from T1 to T3, χ^2^(1) = 40.04, *P*_adjusted_ < .001; and from T2 to T3, χ^2^(1) = 12.07, *P*_adjusted_ < .001. Among women with cannabis use during the first trimester, those reporting sleep as a motive were less likely to discontinue use by the third trimester (26 of 89 participants; 29.2%) than those reporting other motives (49 of 120 participants; 40.5%), although the association was not statistically significant; χ^2^(1) = 3.00, *P* = 0.083.

#### Sleep Outcomes and Cannabis Use During Pregnancy

Main effects of *CUP Status* (0,1) were non-significant for all sleep outcomes (all *P*_FDRs_ <0.59; **eTables 1-4)**. There was a significant main effect of *Trimester* (T1, T2, T3) on sleep quality (F(2,702)=7.54; *P*= 0.001; P_FDR_ =0.002; **eTable 1**) due to lower sleep quality (i.e., higher scores) in the third relative to first and second trimesters (|estimated differences| < 0.17; *Ps* < 0.0001; **Figure 1**; **eTable 5**). No other main effects were observed.

**Figure 1.**
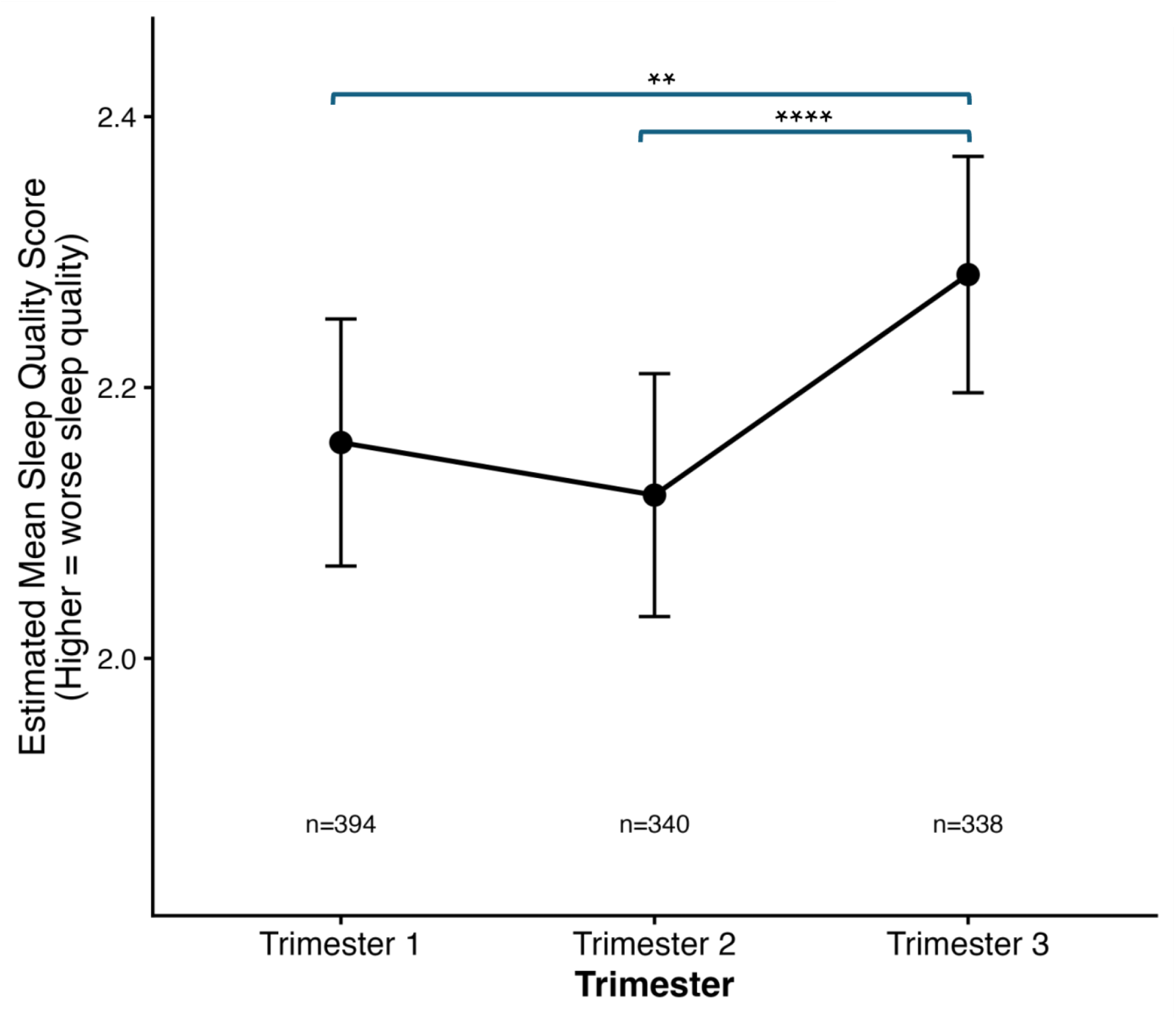
Sleep Quality Was Worse During the Third Trimester. A greater score reflects poorer sleep quality. Lines indicate posthoc contrasts ** = *p* < .01, *** = *p* < .001, **** = *p* < .0001. Data are also available in **eTable 5**.

A *CUP Status* x *Trimester* interaction was associated with insomnia symptoms (F(2,738)=4.98; *P*=0.01, *P*_FDR_ =0.03; **eTable 1**). *Post hoc* contrasts revealed that insomnia symptoms were higher among those using cannabis during the third trimester relative to those who were not. Additionally, there were opposing patterns of insomnia across trimesters by CUP Status; insomnia symptoms were higher in later trimesters (i.e., T3>T2 & T1) among those using cannabis, but lower in later trimesters (i.e., T1>T2 & T3) among those not using cannabis (**Figure 2**; **eFigure 1**; **eTable 6**). The *CUP Status* x *Trimester* interaction and significant *post hoc* comparisons remained significant and were similar in sensitivity analyses covarying for third trimester missingness and when restricted to participants with observed third trimester data (**eTable 3)**. No *CUP Status, Trimester* or *CUP Status* x *Trimester* effects emerged for sleep quality, restless legs symptoms or daytime sleepiness (all Fs<3.22; all *P*_FDRs_ <0.59; **eTables 7-16**).

**Figure 2.**
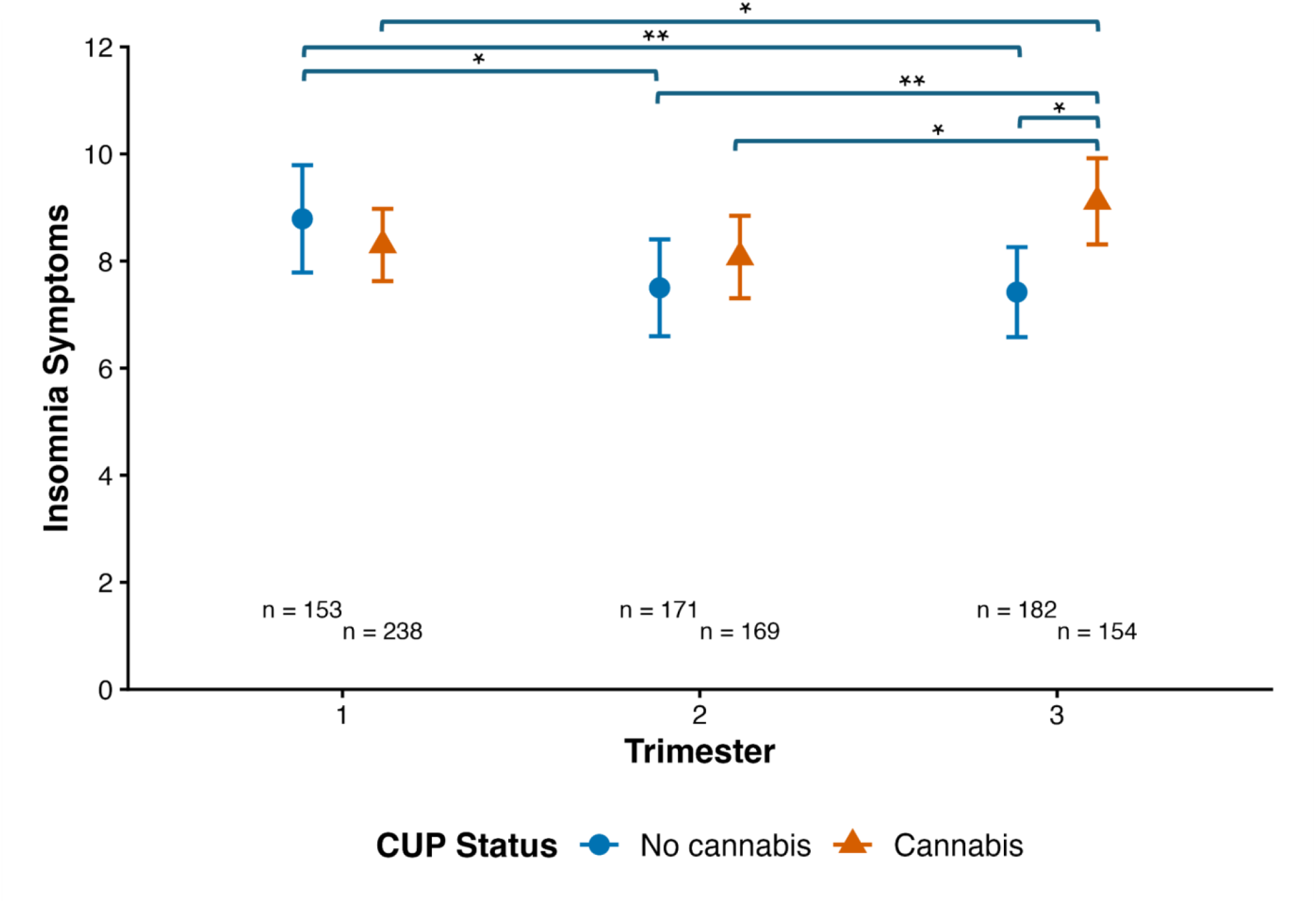
Cannabis Use Status x Trimester Interaction is Associated with Insomnia. CUP Status = cannabis use during pregnancy status at a particular trimester. Insomnia symptoms reported at that trimester assessment. Lines indicate significant *posthoc* contrasts: * = *p* < .05, ** = *p* < .01. Data are also available in **eTable 6**.

#### Frequency and Motives Among Participants Who Used Cannabis During Pregnancy

##### Cannabis Frequency

Among those self-reporting cannabis use (n=286), there were no significant main effects of *Cannabis Use Frequency* nor evidence that *frequency* moderated changes in sleep across trimesters for any sleep phenotype (all Fs < 2.77; all *P*s > 0.11; all *P*_FDRs_>0.39; **eTables 17-20**). ***Motives***. Reporting sleep as the primary motive for CUP in the first trimester was associated with more insomnia symptoms and worse sleep quality across pregnancy (Fs> 7.85, *Ps* < 0.005, P_FDRs_<0.006; **Figure 3, eTables 21**). Sleep motive and its interaction with trimester were not associated with any other sleep outcome (Fs<3.16; all *P*s > 0.045; *P*_FDRs_<0.18; **eTable 21-26**).

**Figure 3.**
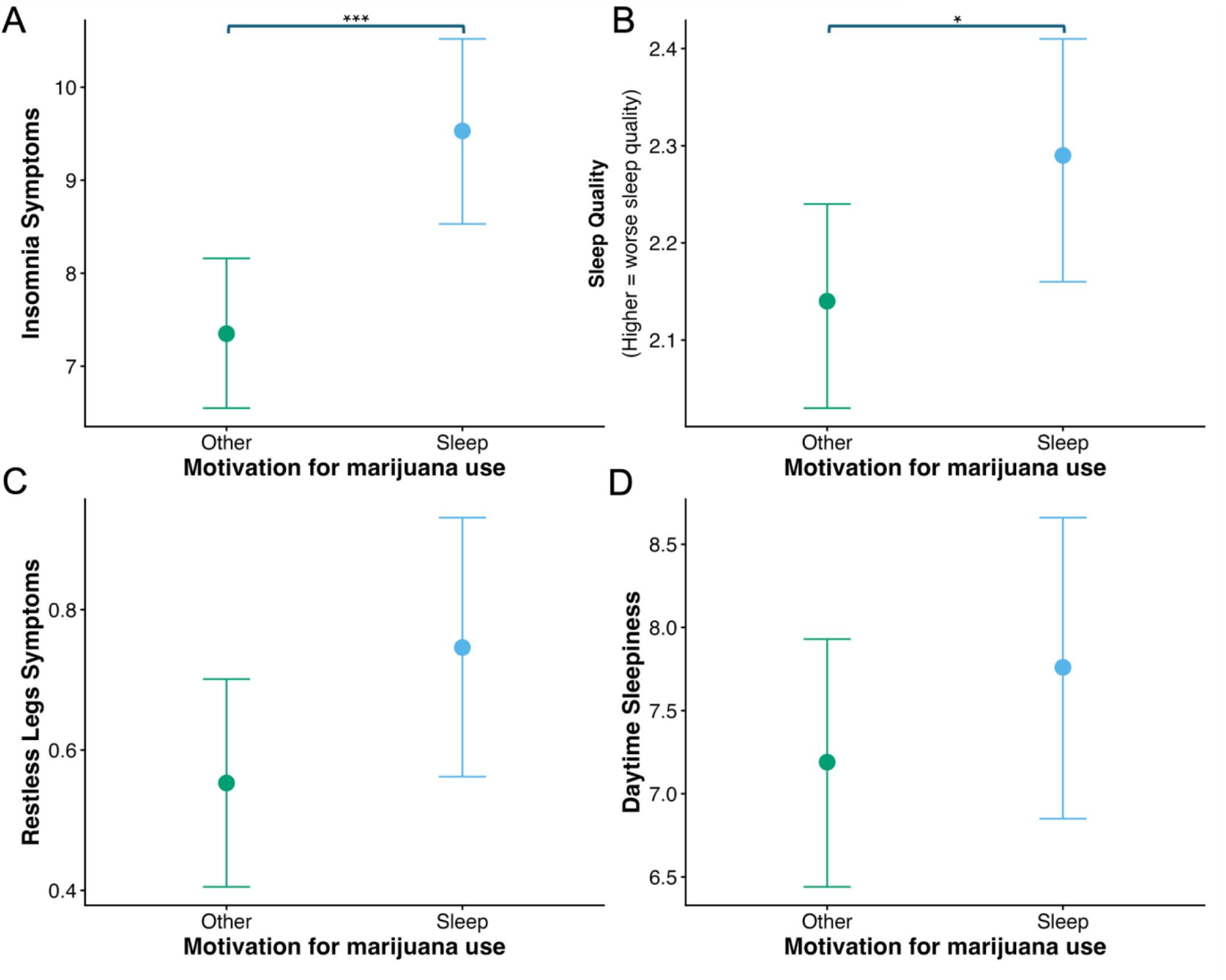
Sleep Motives for Cannabis Use during Pregnancy and Sleep Problems. Reporting cannabis use during pregnancy to address sleep problems was associated with significantly higher insomnia **(A)** and worse sleep quality **(B)**, but no differences in restless legs symptoms **(C)** or daytime sleepiness **(D)** Asterisks denote statistical significance. * = *p* < .05, *** = *p* < .001.

## DISCUSSION

Cannabis use during pregnancy (CUP) remains common, and sleep problems are among the most frequently cited motivations for use.^7^ However, evidence regarding the association between cannabis use and sleep remains mixed in general population samples^12,13,16^ and randomized clinical trials^15^ with few studies examining this association in pregnant populations.^20,21^ Here, we found evidence that cannabis use was associated with worse insomnia symptoms, but not sleep quality, restless legs, of daytime sleepiness during later pregnancy. Moreover, women who reported using cannabis to help sleep experienced worse insomnia symptoms and poorer sleep quality than those who used cannabis for other reasons. As sleep problems are a common motive for CUP and prenatal exposure to cannabis and maternal sleep problems are associated with adverse outcomes, increasing knowledge of and access to empirically supported options to promote healthy sleep during pregnancy (e.g., cognitive behavioral therapy for insomnia)^35^ may benefit the health of mothers and offspring through attenuated cannabis use and exposure to maternal sleep problems during pregnancy.

Our findings extend a limited pregnancy-specific literature^20,21^ by examining a larger sample and modeling CUP separately within each trimester, in contrast to prior studies that characterized CUP more broadly.^20,21^ This approach identified trimester-specific differences where women who used cannabis reported greater insomnia symptom severity in the third trimester compared to other trimesters. As CUP status was modeled to vary across trimesters, these findings should be interpreted as trimester-specific associations rather than comparisons of fixed cannabis use groups. Although insomnia typically worsens as pregnancy progresses,^5^ women who did not use cannabis in the current cohort had lower insomnia symptoms in later pregnancy than in the first trimester. This pattern suggests that cannabis use in the third trimester may identify women experiencing a particularly high burden of insomnia during a period when sleep problems are already common.

Several non-mutually exclusive factors may contribute to the association between third trimester cannabis use and greater insomnia symptom severity (e.g., shared predispositional risk, shared social or behavioral risk factors).^1,8,16,36,37^ Women who used in the third trimester may represent a time-varying subgroup, with persistent or recurrent use, greater social or structural vulnerability, and/or fewer alternatives for managing sleep problems. Consistent with this notion, women who reported sleep as a motive for CUP had greater insomnia symptoms and poorer sleep quality than those who reported other motives, paralleling prior findings that sleep-motivated CUP is associated with greater sleep problems.^21^ Frequency was not associated with any sleep outcomes. Although these findings do not suggest a frequency-response relationship, frequency alone may not fully capture variation in dose, potency, timing, product-type, or route of administration.^38,39^ Neither cotinine status nor its interaction with CUP explained the association between CUP and third-trimester insomnia, suggesting that nicotine exposure alone was unlikely to account for this finding.^40^

Cannabis use was not associated with overall sleep quality, consistent with prior work in pregnant women^20^ and randomized clinical trials,^15^ although findings in general populations have been mixed.^11–13,16,19,41^ We also found no association between CUP and daytime sleepiness or restless legs symptom severity, which have yet to be examined in prior literature to our knowledge. Emerging evidence in the general population suggests that associations between use and sleep may differ by measurement method, with studies of subjective sleep reporting mixed findings,^14,16,19,41^ whereas studies using objective sleep measures have generally observed increased sleep problems.^12,14,42,43^ This may arise from subjective reports being influenced by perceptions that cannabis improves sleep^9^ as well as equivocal evidence from administration studies that cannabis may impact unobservable sleep architecture (e.g., suppressing rapid eye movement sleep) while not influencing more readily observable (e.g., total sleep time, sleep latency, wake time) aspects of sleep that may influence subjective reports.^15^ Future research using both objective and subjective sleep measures is needed to better characterize the association between CUP and sleep. Collectively, these findings suggest that insomnia symptom severity may be an important clinical context for understanding CUP, particularly among women who reported using it to improve sleep as their motivation for use.

### Limitations

Several limitations should be considered when interpreting these findings. *First*, our study was composed of predominantly under-resourced individuals (income-to-needs ratio average: 1.21±0.99; Range: 0.34-9.72) who may experience elevated structural stressors,^44^ environmental exposures,^45^ and reduced access to sleep-promoting resources,^44^ which may contribute to sleep experiences within participants in our study relative to the general population. ^46^ Critically, however, modeling income-to-needs differences across cannabis use groups did not modify the significance of observed associations. *Second*, sleep was assessed using self-report, which may introduce subjective biases and fail to capture sleep problems as accurately as objective measures, particularly given recent evidence of associations between cannabis use and objectively, but not subjectively, measured sleep problems.^12,47,48^ *Third*, as the study was not specifically designed to study sleep, variability in the daily timing of cannabis use, which was not assessed, may have impacted our findings; for example, it is possible that those reporting cannabis use for sleep are more likely to use cannabis before attempting to sleep. Future clinical trials are needed to clarify the temporal relationship and potential causal nature of these associations. *Fourth*, sleep aids (e.g., non-reported melatonin use) other than medications available from the medical record were not evaluated and may have impacted sleep and introduced variability into association estimates. *Fifth*, a small portion of data was obtained retrospectively when a trimester clinical care assessment was missed which may introduce inaccuracies of report. However, sensitivity analyses including an indicator for participants with first-trimester data but missing-third trimester cannabis and/or sleep outcome data did not meaningfully change current findings.

## Conclusion

In this cohort study of pregnant women, cannabis use in the third trimester was associated with greater insomnia symptom severity. Further, women who reported using cannabis to manage sleep problems had greater insomnia symptom severity and poorer sleep quality than those who used cannabis for other reasons. These findings suggest that insomnia may be an important clinical context for understanding cannabis use during pregnancy, particularly when the motivation for use is to manage sleep. Improving access to evidence-based approaches for managing sleep during pregnancy may therefore be of particular importance as a component of prenatal care and warrants further evaluation as potential strategy for reducing sleep-motivated prenatal cannabis use.

## Acknowledgements

We are thankful to families who have participated in the CUDDEL Study as well as study staff and investigators.

## Author Contributions

Gorelik and Drs Agrawal and Johnson had full access to all of the data in the study and take responsibility for the integrity of the data and the accuracy of the data analysis.

### Concept and design

Gorelik, Bogdan, Agrawal, Johnson

### Acquisition, analysis, or interpretation of data

Gorelik, Cunado, Kinstler, Lenze, Smyser, Rogers, Bogdan, Agrawal, Johnson

### Drafting and editing of the manuscript

Gorelik, Cunado, Kinstler, Bogdan, Agrawal, Johnson, Cox

### Critical review of the manuscript for important intellectual content

Gorelik, Cunado, Kinstler, Constantino-Petti, Leverett, Lenze, Smyser, Rogers, Bogdan, Agrawal, Johnson, Cox

### Statistical analysis

Gorelik, Johnson

### Obtained funding

Johnson, Bogdan, Rogers, Agrawal

### Administrative, technical, or material support

Agrawal, Bogdan.

### Supervision

Rogers, Bogdan, Agrawal, Johnson

## Declarations

### Data availability

CUDDEL data will be made available through the National Data Archive

### Code

https://github.com/WashU-BG/maternal_sleep_during_pregnancy_cannabis/tree/analyses-code

