## Supplement for "Associations Between Cannabis Use and Sleep Outcomes in a Sample of Pregnant Women"

This supplemental material has been provided by the authors to give readers additional information about the current work.

### eMethods

**CUDDEL Sample.** The ongoing Cannabis Use During Early Life and Development (CUDDEL) study is a longitudinal cohort study that recruited 461 pregnant individuals who used and did not use cannabis during pregnancy to study correlates of use among mothers and exposure among offspring.[1] Pregnant participants were recruited from an obstetric clinic in an academic hospital between July 2019 and January 2024. At enrollment, all participants reported a lifetime history of cannabis use prior to their current pregnancy. Inclusion criteria included: 1) lifetime history of cannabis use prior to pregnancy (even among those not using cannabis during pregnancy) 2) ≥18 years of age, 3) English speaking, 4) ability to provide informed consent, 5) singleton pregnancy, 6) lack of current incarceration, 7) no use of reproductive technology in conception, 8) report of no tobacco or other illicit drug use during pregnancy, 9) negative urine drug testing results for all substances other than cannabis at each assessment, with the exception of positivity for the nicotine within the cannabis use group, due to common blunt usage, 10) report of no heavy episodic alcohol consumption following knowledge of pregnancy, 11) report of no use of or prescriptions for prescription medications with known serious effects on fetal brain development, 12) no diagnosis of active psychosis, mania, or suicidal ideation. Exclusion criteria included: 1) incarceration; 2) age younger than 18 years at delivery; 3) active psychosis, mania, or suicidal ideation; 4) multifetal pregnancy; 5) use of assisted reproductive therapy; 6) self-report of heavy alcohol use or any illicit or potentially teratogenic substance use following knowledge of pregnancy including tobacco, cocaine (crack, rock, ice), methamphetamine (speed, crank), heroin, or opiates (when not as prescribed), hallucinogens, PCP, Steroids, MDMA; and 7) urine positivity during any point in pregnancy for (see ***Other Substance Use and Cotinine Positivity*** below). All participants provided written informed consent to a protocol approved by the Washington University institutional review board. This study followed the Strengthening the Reporting of Observational Studies in Epidemiology (STROBE)[2] reporting guidelines.

### Measures

**Cannabis Use During Pregnancy.** Trimester-specific cannabis use during pregnancy (CUP) status, which could vary within individuals, was coded as positive for participants self-reporting cannabis use *or* having urine positivity during study visits ($n_{T1}$=261 [60.6%], $n_{T2}$=212 [51.1%], $n_{T3}$=178 [44.7%]) and negative when participants reported no use *and* tested negative in available urine samples at a given trimester visit ($n_{T1}$=170 [39.4%], $n_{T2}$=203 [48.9%], $n_{T3}$=220 [55.3%]). Binary CUP Status (cannabis use versus no use) had moderate stability across trimesters: T1-T2 r=.601, T1-T3 r=.521, T2-T3 r=.696. ***Self-report.*** Participants reported their frequency of use (i.e., never, monthly or less, 2-4x/month, 2-3x/week) at each trimester. This question was based on item 1 of the Cannabis Use Disorders Identification Test-Revised (CUDIT-R),[3] with modifications to assess use during pregnancy during the first trimester prior to and subsequent to knowledge of pregnancy and since the prior study visit in trimesters 2 and 3. The following frequencies were reported: never ($n_{T1}$=186, $n_{T2}$=191, $n_{T3}$=218), monthly or less ($n_{T1}$=68, $n_{T2}$=45, $n_{T3}$=41), 2 to 4 times a month ($n_{T1}$=38, $n_{T2}$=40, $n_{T3}$=30), 2 to 3 times a week ($n_{T1}$=47, $n_{T2}$=36, $n_{T3}$=27), or 4 or more times a week ($n_{T1}$=33, $n_{T2}$=27, $n_{T3}$=25). For analyses, cannabis frequency was recoded at each trimester as never ($n_{T1}$=152, $n_{T2}$=168, $n_{T3}$=191), monthly ($n_{T1}$=68, $n_{T2}$=45, $n_{T3}$=41), or at least 2 or more times per month ($n_{T1}$=118, $n_{T2}$=103, $n_{T3}$=82). ***Urine Drug Testing.*** At each clinical care trimester visit urine was tested for the cannabis metabolite THC-COOH (i.e., 11-nor-Δ9-THC-9 COOH; cutoff concentration of 50ng/mL; estimated detection window of approximately 3-30 days) using commercially available urine drug testing kits (i.e., Test Country, 12 panel cup; Confirm Biosciences 12 panel cup/DrugConfirm; or 12panelnow 10 panel cup). Concordance between self-reported cannabis use and THC positive urine screens was moderate (r = 0.7), indicating reasonable agreement.[1]

***Cannabis Use Motives.*** Among women reporting cannabis use during pregnancy, motives for use were assessed during the first trimester with the following prompt, "*Different women have different reasons for using marijuana during pregnancy. Please mark all that apply to you.*" Listed motives for participant selection (multiple response options permitted) were drawn from the *Pregnancy Risk Assessment Monitoring Survey*[5] and other measures and included: 1) to deal with nausea/vomiting, 2) to deal with anxiety or stress, 3) to help with hunger or loss of appetite, 4) to help you sleep, 5) to treat mental health problems (e.g., depression, posttraumatic stress disorder), 6) for enjoyment and relaxation, 7) to have energy, get more things done during the day, 8) to help with pain, 9) it was uncomfortable to stop using, and 10) other (with write-in allowed).

***Other Substance Use and Cotinine Positivity.*** Self-report of other substance use or urine positivity at any point during pregnancy was exclusionary. Study eligibility included lack of tobacco use; however, a proportion of participants reported using cannabis in a manner that typically involves tobacco products as well as second hand exposure. As a result, participants who used cannabis during pregnancy and tested positive for cotinine were not excluded from the study. Within the group of individuals reporting cannabis use or testing positive for THC in urine (n = 298 across trimesters), 158 tested positive for cotinine. As such cotinine across pregnancy was included as a fixed effect to account for this. ***Urine Drug Testing***. At each trimester clinical care visit, urine was tested for the presence of other substance metabolites (see above), using commercially available drug testing cups((Test Country, 12 panel cup; Confirm Biosciences 12 panel cup/DrugConfirm; 12panelnow 10 panel cup). Participants were excluded from the study if they reported use of these substances or tested positive for any of these substances at any trimester assessment. The nicotine metabolite cotinine was tested for in urine at each trimester visit using a commercially available cotinine strip test (Premier Brands; concentration cut off: 200ng/mL; estimated detection window of up to 10 days). Women who used cannabis and reported no nicotine use, but tested positive for cotinine, were retained within the study as this may be attributable to blunts as a mode of cannabis use in this sample and/or second-hand exposure. Among participants who reported cannabis use or tested positive for cannabis during pregnancy, 158 (37.2%) tested positive for cotinine and 267 (62.8%) tested negative. Ever testing positive at any trimester was used as a covariate in analyses. Women without evidence of cannabis use at any trimester visit who tested positive for cotinine were excluded from the study.

**Sleep Indices.** Sleep phenotypes across trimesters included insomnia symptoms (Women's Health Initiative Insomnia Rating Scale)[6], daytime sleepiness (Epworth Sleepiness Scale),[7] restless legs symptoms, (International Restless Legs Syndrome Study Group Severity Scale),[8], and subjective sleep quality (global item of the Pittsburgh Sleep Quality Index).[9] See below for more details:

Women's Health Initiative Insomnia Rating Scale (WHIIRS): Responses were scored on a Likert scale ranging from 0 (no difficulty) to 4 (very frequent difficulty), with total scores ranging from 0 to 20. Higher scores indicated greater severity of insomnia symptoms.

Epworth Sleepiness Scale (ESS): Participants rated their likelihood of dozing off in various situations, such as sitting and reading or watching television, on a scale from 0 (no chance of dozing) to 3 (high chance of dozing). Total scores range from 0 to 24, with higher scores indicating higher levels of daytime sleepiness.

Restless Legs Symptom (RLS): to evaluate features of RLS, including an urge to move the legs, worsening of symptoms during rest or inactivity, improvement with movement, and diurnal variation, with symptoms predominantly occurring in the evening or night.

Sleep Quality (PSQI). Completion of the full Pittsburgh Sleep Quality Index (PSQI) varied across trimesters, resulting in substantial missingness. Accordingly, we operationalized subjective sleep quality using the single-item PSQI global sleep quality question, which was available in most of participants.[9] Participants were asked, "During the past month, how would you rate your sleep quality overall?" with response options of very good, fairly good, fairly bad, or very bad.[9]

**Participant Race & Ethnicity**

Participants were asked to self-identify their race (i.e., American Indian or Alaska Native, Asian, Black or African American, Hawaiian or Pacific Islander, White, and "other"; multiple category responses permitted) and ethnicity (i.e., Hispanic or Latinx, non-Hispanic or Latinx, and unknown) during their first trimester to characterize the racial and ethnic make-up of the sample.

**Income-to-needs Ratios**

Income-to-needs ratios were derived by dividing participant reported total household income (assessed during the first trimester) by the federal guideline for poverty (from the same year the participant reported), given participant reported family size. Lower income-to-needs ratios denote greater socioeconomic disadvantage.

**eFigure 1. Pairwise Contrasts of Adjusted Insomnia Scores Across Trimester x Cannabis Use During Pregnancy Status.** Estimated marginal means for each trimester x cannabis group status from the mixed-effects model and compared in 36 pairwise contrasts. Points reflect the estimated difference in adjusted mean insomnia scores for each contrast. Error bars represent 95% CIs and orange points signify p<0.05.

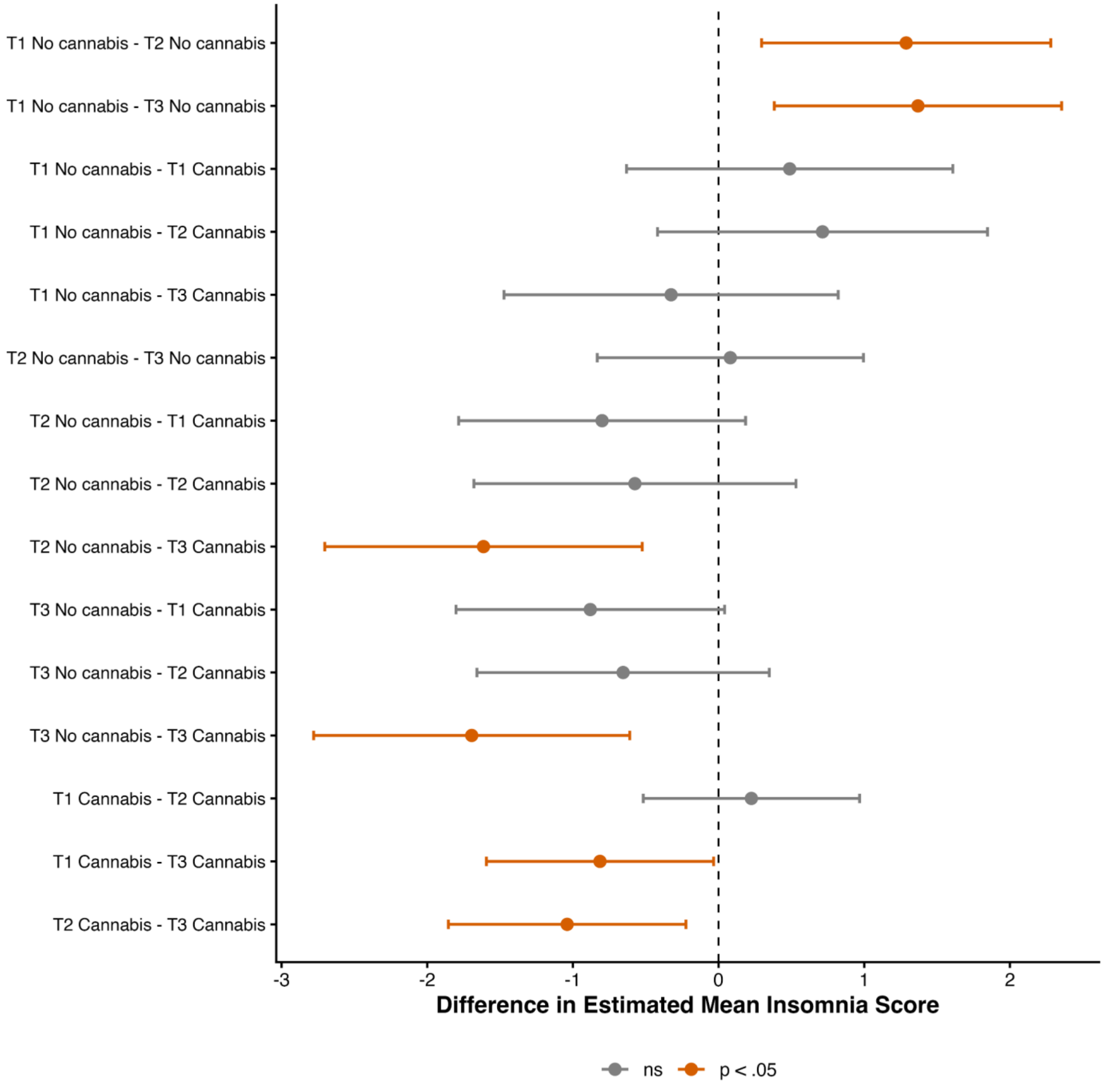

**eTables 1-4 Primary Analyses:** Maternal Sleep Across Trimesters**.** False discovery rate correction was conducted for each respective variable across all four sleep phenotypes

**eTable 1:** Maternal Restless Legs Symptoms Across Trimester

| Variable | Sum Sq | Mean Sq | NumDF | DenDF | F value | p-value | p_FDR |
|---|---|---|---|---|---|---|---|
| trimester | 3.29 | 1.65 | 2 | 779 | 2.8 | 0.061 | 0.065 |
| CUP Status | 0.92 | 0.92 | 1 | 1142 | 1.57 | 0.211 | 0.281 |
| age | 4.8 | 4.8 | 1 | 431 | 8.17 | 0.005 | - |
| income to needs ratio | 1.69 | 1.69 | 1 | 426 | 2.87 | 0.091 | - |
| cotinine status | 0.16 | 0.16 | 1 | 493 | 0.27 | 0.602 | 0.602 |
| trimester × CUP Status | 2.9 | 1.45 | 2 | 869 | 2.47 | 0.086 | 0.171 |
| trimester × age | 1.55 | 0.77 | 2 | 779 | 1.32 | 0.268 | - |
| trimester × income to needs ratio | 14.07 | 7.04 | 2 | 766 | 11.97 | 7.60E-06 | - |
| trimester × cotinine status | 2.72 | 1.36 | 2 | 807 | 2.31 | 0.100 | - |
| CUP Status × age | 0.02 | 0.02 | 1 | 1096 | 0.04 | 0.844 | - |
| CUP Status × income to needs ratio | 0.17 | 0.17 | 1 | 974 | 0.29 | 0.589 | - |
| CUP Status × cotinine status | 0.02 | 0.02 | 1 | 1137 | 0.03 | 0.862 | - |

**eTable 2:** Maternal Insomnia Symptoms Across Trimesters

| Variable | Sum Sq | Mean Sq | NumDF | DenDF | F value | p-value | p_FDR |
|---|---|---|---|---|---|---|---|
| trimester | 66.15 | 33.07 | 2 | 686.66 | 2.75 | 0.06 | 0.06 |
| CUP Status | 30.78 | 30.78 | 1 | 1047.76 | 2.56 | 0.11 | 0.28 |
| age | 57.64 | 57.64 | 1 | 403.22 | 4.79 | 0.03 | - |
| income to needs ratio | 1.54 | 1.54 | 1 | 399.31 | 0.13 | 0.72 | - |
| cotinine status | 8.25 | 8.25 | 1 | 462.42 | 0.69 | 0.41 | - |
| **trimester × CUP Status** | **119.81** | **59.91** | **2** | **737.81** | **4.98** | **0.01** | **0.03** |
| trimester × age | 27.55 | 13.78 | 2 | 679.94 | 1.15 | 0.32 | - |
| trimester × income to needs ratio | 47.89 | 23.94 | 2 | 668.09 | 1.99 | 0.14 | - |
| trimester × cotinine status | 44.87 | 22.43 | 2 | 702.66 | 1.87 | 0.16 | 0.31 |
| CUP Status × age | 1.25 | 1.25 | 1 | 1048.42 | 0.10 | 0.75 | - |
| CUP Status × income to needs ratio | 4.44E-04 | 4.44E-04 | 1 | 971.59 | 0.00 | 1.00 | - |
| CUP Status × cotinine status | 0.741 | 0.741 | 1 | 986.08 | 0.062 | 0.804 | - |

**eTable 3:** Maternal Daytime sleepiness Across Trimesters

| Variable | Sum Sq | Mean Sq | NumDF | DenDF | F value | p-value | p_FDR |
|---|---|---|---|---|---|---|---|
| trimester | 73.2 | 36.6 | 2 | 602 | 3.21 | 0.041 | 0.065 |
| CUP Status | 3.5 | 3.5 | 1 | 936 | 0.31 | 0.580 | 0.580 |

| | | | | | | | |
|---|---|---|---|---|---|---|---|
| age | 50.1 | 50.1 | 1 | 397 | 4.38 | 0.037 | - |
| income to needs ratio | 1.4 | 1.4 | 1 | 375 | 0.13 | 0.723 | - |
| cotinine status | 54.7 | 54.7 | 1 | 451 | 4.79 | 0.029 | - |
| trimester × CUP Status | 23.9 | 11.9 | 2 | 660 | 1.04 | 0.353 | - |
| trimester × age | 89.2 | 44.6 | 2 | 595 | 3.9 | 0.021 | - |
| trimester × income to needs ratio | 19.1 | 9.6 | 2 | 568 | 0.84 | 0.434 | - |
| trimester × cotinine status | 2.2 | 1.1 | 2 | 618 | 0.1 | 0.908 | 0.908 |
| CUP Status × age | 10.9 | 10.9 | 1 | 931 | 0.95 | 0.329 | - |
| CUP Status × income to needs ratio | 0 | 0 | 1 | 807 | 0 | 0.991 | - |
| CUP Status × cotinine status | 2 | 2 | 1 | 906 | 0.17 | 0.677 | - |

**eTable 4:** Maternal Sleep Quality Across Trimesters

| Variable | Sum Sq | Mean Sq | NumDF | DenDF | F value | p-value | p_FDR |
|---|---|---|---|---|---|---|---|
| **trimester** | **4.72** | **2.36** | **2** | **702** | **7.54** | **0.001** | **0.002** |
| CUP Status | 0.69 | 0.688 | 1 | 1024 | 2.2 | 0.138 | 0.277 |
| age | 1.22 | 1.221 | 1 | 397 | 3.9 | 0.049 | - |
| income to needs ratio | 0 | 0 | 1 | 391 | 0 | 0.981 | - |
| cotinine status | 0.1 | 0.097 | 1 | 460 | 0.31 | 0.578 | - |
| trimester × CUP Status | 0.43 | 0.214 | 2 | 783 | 0.68 | 0.505 | 0.505 |
| trimester × age | 2.42 | 1.21 | 2 | 693 | 3.87 | 0.021 | - |
| trimester × income to needs ratio | 0.87 | 0.436 | 2 | 679 | 1.39 | 0.249 | - |
| trimester × cotinine status | 0.07 | 0.035 | 2 | 727 | 0.11 | 0.895 | 0.908 |
| CUP Status × age | 0.21 | 0.209 | 1 | 992 | 0.67 | 0.413 | - |
| CUP Status × income to needs ratio | 0.13 | 0.127 | 1 | 841 | 0.41 | 0.524 | - |
| CUP Status × cotinine status | 0.12 | 0.117 | 1 | 1048 | 0.38 | 0.540 | - |

**eTable 5:** Pairwise Contrasts for Maternal Sleep Quality Across Trimesters

| Result type | Trimester/ Contrast | Emmean /Estimate | SE | df | Lower CL | Upper CL | t ratio | p-value |
|---|---|---|---|---|---|---|---|---|
| Estimated mean | Trimester 1 | 2.16 | 0.0465 | 960 | 2.07 | 2.25 | - | - |
| Estimated mean | Trimester 2 | 2.12 | 0.0457 | 951 | 2.03 | 2.21 | - | - |
| Estimated mean | Trimester 3 | 2.28 | 0.0445 | 931 | 2.2 | 2.37 | - | - |
| Contrast | Trimester 1-Trimester 2 | 0.0388 | 0.0466 | 714 | - | - | 0.83 | 0.405 |
| Contrast | **Trimester 1-Trimester 3** | **-0.124** | **0.0464** | **723** | - | - | **-2.67** | **0.008** |

| | | | | | | | | | |
|---|---|---|---|---|---|---|---|---|---|
| Contrast | **Trimester 2-Trimester 3** | | **-0.1628** | **0.0464** | **696** | - | - | **-3.51** | **<.0001** |

**eTable 6:** Pairwise contrasts for Maternal insomnia Symptoms Across Trimesters

| Result type | Trimester/ Contrast | CUP Status | Emmean /Estimate | SE | df | Lower CL | Upper CL | t ratio | p-value |
|---|---|---|---|---|---|---|---|---|---|
| Estimated mean | Trimester 1 | nocan | 8.79 | 0.511 | 1037 | 7.79 | 9.79 | - | - |
| Estimated mean | Trimester 2 | nocan | 7.5 | 0.461 | 1047 | 6.6 | 8.41 | - | - |
| Estimated mean | Trimester 3 | nocan | 7.42 | 0.428 | 1048 | 6.58 | 8.26 | - | - |
| Estimated mean | Trimester 1 | can | 8.3 | 0.344 | 891 | 7.63 | 8.98 | - | - |
| Estimated mean | Trimester 2 | can | 8.08 | 0.392 | 991 | 7.31 | 8.85 | - | - |
| Estimated mean | Trimester 3 | can | 9.11 | 0.41 | 1015 | 8.31 | 9.92 | - | - |
| Contrast | **Trimester 1 nocan-Trimester 2 nocan** | - | **1.288** | **0.506** | **733** | - | - | **2.547** | **0.011** |
| Contrast | **Trimester 1 nocan-Trimester 3 nocan** | - | **1.369** | **0.502** | **740** | - | - | **2.725** | **0.007** |
| Contrast | Trimester 1 nocan-Trimester 1 can | - | 0.488 | 0.571 | 881 | - | - | 0.856 | 0.392 |
| Contrast | Trimester 1 nocan-Trimester 2 can | - | 0.714 | 0.577 | 888 | - | - | 1.236 | 0.217 |
| Contrast | Trimester 1 nocan-Trimester 3 can | - | -0.326 | 0.585 | 895 | - | - | -0.558 | 0.577 |
| Contrast | Trimester 2 nocan-Trimester 3 nocan | - | 0.081 | 0.465 | 699 | - | - | 0.174 | 0.862 |
| Contrast | Trimester 2 nocan-Trimester 1 can | - | -0.8 | 0.502 | 864 | - | - | -1.593 | 0.111 |
| Contrast | Trimester 2 nocan-Trimester 2 can | - | -0.574 | 0.563 | 885 | - | - | -1.02 | 0.308 |
| Contrast | **Trimester 2 nocan-Trimester 3 can** | - | **-1.614** | **0.555** | **901** | - | - | **-2.907** | **0.004** |
| Contrast | Trimester 3 nocan-Trimester 1 can | - | -0.881 | 0.47 | 857 | - | - | -1.875 | 0.061 |
| Contrast | Trimester 3 nocan-Trimester 2 can | - | -0.655 | 0.511 | 884 | - | - | -1.282 | 0.2 |
| Contrast | **Trimester 3 nocan-Trimester 3 can** | - | **-1.695** | **0.553** | **887** | - | - | **-3.065** | **0.002** |
| Contrast | Trimester 1 can-Trimester 2 can | - | 0.225 | 0.378 | 713 | - | - | 0.596 | 0.552 |
| Contrast | **Trimester 1 can-Trimester 3 can** | - | **-0.814** | **0.397** | **729** | - | - | **-2.05** | **0.041** |
| Contrast | **Trimester 2 can-Trimester 3 can** | - | **-1.039** | **0.415** | **690** | - | - | **-2.503** | **0.013** |

**eTables 7-11** Sensitivity Analyses.

| Effect | Sum Sq | Mean Sq | NumDF | DenDF | F value | p-value |
|---|---|---|---|---|---|---|
| **eTable 7:** Insomnia Symptom Analyses Adjusted for Missing T3 Insomnia Indicator | | | | | | |
| trimester | 66.1 | 33 | 2 | 674 | 2.75 | 0.0648 |
| CUP Status | 30.4 | 30.4 | 1 | 1047 | 2.53 | 0.1121 |
| t1_cup_t3_cup_missing | 1.7 | 1.7 | 1 | 514 | 0.14 | 0.7097 |
| **age** | **57.5** | **57.5** | **1** | **402** | **4.78** | **0.0293** |
| income to need ratio | 1.5 | 1.5 | 1 | 398 | 0.12 | 0.7253 |
| cotinine status | 8.3 | 8.3 | 1 | 461 | 0.69 | 0.4069 |
| **Trimester * CUP Status** | **119.9** | **59.9** | **2** | **737** | **4.98** | **0.0071** |
| trimester * age | 27.5 | 13.8 | 2 | 680 | 1.14 | 0.3191 |
| trimester * cotinine status | 44.7 | 22.4 | 2 | 702 | 1.86 | 0.1564 |
| trimester * income to need ratio | 48.1 | 24 | 2 | 668 | 2 | 0.1364 |
| CUP Status * age | 1.3 | 1.3 | 1 | 1047 | 0.11 | 0.7444 |
| CUP Status * income to need ratio | 0 | 0 | 1 | 970 | 0 | 0.9886 |
| CUP Status * cotinine status | 0.8 | 0.8 | 1 | 985 | 0.06 | 0.8027 |
| **eTable 8:** Insomnia Symptom Analyses Adjusted for Missing T3 CUP Status Indicator | | | | | | |
| trimester | 66.2 | 33.1 | 2 | 681 | 2.75 | 0.065 |
| CUP Status | 30 | 30 | 1 | 1047 | 2.49 | 0.115 |
| t1_cup_t3_cup_missing | 4.4 | 4.4 | 1 | 517 | 0.36 | 0.548 |
| **age** | **57.2** | **57.2** | **1** | **402** | **4.76** | **0.03** |
| income to need ratio | 1.4 | 1.4 | 1 | 398 | 0.11 | 0.736 |
| cotinine status | 8.1 | 8.1 | 1 | 461 | 0.68 | 0.411 |
| **trimester * CUP Status** | **120.1** | **60.1** | **2** | **737** | **4.99** | **0.007** |
| trimester * age | 27.4 | 13.7 | 2 | 679 | 1.14 | 0.32 |
| trimester * cotinine status | 44.9 | 22.4 | 2 | 702 | 1.87 | 0.155 |
| trimester * income to need ratio | 48 | 24 | 2 | 667 | 2 | 0.137 |
| CUP Status * age | 1.2 | 1.2 | 1 | 1047 | 0.1 | 0.755 |
| CUP Status * income to need ratio | 0 | 0 | 1 | 970 | 0 | 0.994 |
| CUP Status * cotinine status | 0.8 | 0.8 | 1 | 985 | 0.07 | 0.798 |
| **eTable 9:** Insomnia Analyses Adjusted for Both T3 Missingness Indicators | | | | | | |
| trimester | 66.1 | 33.1 | 2 | 674 | 2.75 | 0.065 |
| CUP Status | 30 | 30 | 1 | 1046 | 2.5 | 0.114 |
| t1_cup_t3_cup_missing | 2.7 | 2.7 | 1 | 521 | 0.22 | 0.637 |
| t1_whiirs_t3_whiirs_missing | 0 | 0 | 1 | 518 | 0 | 0.985 |
| **age** | **57.1** | **57.1** | **1** | **401** | **4.75** | **0.03** |
| income to need ratio | 1.4 | 1.4 | 1 | 398 | 0.11 | 0.736 |
| cotinine status | 8.1 | 8.1 | 1 | 460 | 0.68 | 0.412 |
| **trimester * CUP Status** | **120.1** | **60** | **2** | **736** | **4.99** | **0.007** |
| trimester * age | 48 | 24 | 2 | 667 | 1.99 | 0.137 |
| trimester * cotinine status | 27.4 | 13.7 | 2 | 679 | 1.14 | 0.32 |
| trimester * income to need ratio | 44.9 | 22.4 | 2 | 701 | 1.87 | 0.156 |
| CUP Status * age | 1.2 | 1.2 | 1 | 1046 | 0.1 | 0.755 |
| CUP Status * income to need ratio | 0 | 0 | 1 | 969 | 0 | 0.994 |
| CUP Status * cotinine status | 0.8 | 0.8 | 1 | 984 | 0.07 | 0.798 |
| **eTable 10:** Insomnia Analyses Restricted to Observed T3 CUP Status and Insomnia Symptoms | | | | | | |
| trimester | 33.8 | 16.9 | 2 | 622 | 1.37 | 0.2545 |
| CUP Status | 32.8 | 32.8 | 1 | 935 | 2.66 | 0.103 |

| | | | | | | |
|---|---|---|---|---|---|---|
| **age** | **83.9** | **83.9** | **1** | **335** | **6.81** | **0.0095** |
| income to need ratio | 0.2 | 0.2 | 1 | 336 | 0.02 | 0.8925 |
| cotinine status | 3.6 | 3.6 | 1 | 382 | 0.29 | 0.5894 |
| **trimester * CUP Status** | **125** | **62.5** | **2** | **662** | **5.07** | **0.0065** |
| trimester * age | 17.7 | 8.9 | 2 | 621 | 0.72 | 0.4876 |
| trimester * cotinine status | 53 | 26.5 | 2 | 636 | 2.15 | 0.1173 |
| trimester * income to need ratio | 46.7 | 23.3 | 2 | 614 | 1.89 | 0.1515 |
| CUP Status * age | 0 | 0 | 1 | 931 | 0 | 0.9802 |
| CUP Status * income to need ratio | 1.3 | 1.3 | 1 | 848 | 0.1 | 0.7483 |
| CUP Status * cotinine status | 0.8 | 0.8 | 1 | 891 | 0.06 | 0.8005 |

**eTable 11:** Insomnia Analyses Restricted to Cannabis Use During Pregnancy Status Data Available at All Three Trimesters

| | | | | | | |
|---|---|---|---|---|---|---|
| trimester | 43.1 | 21.5 | 2 | 622 | 1.79 | 0.1679 |
| CUP Status | 21.9 | 21.9 | 1 | 938 | 1.82 | 0.1776 |
| **age** | **56.2** | **56.2** | **1** | **336** | **4.67** | **0.0315** |
| income to needs ratio | 1.9 | 1.9 | 1 | 337 | 0.15 | 0.6947 |
| cotinine status | 7.5 | 7.5 | 1 | 382 | 0.62 | 0.4317 |
| **trimester * CUP Status** | **132.8** | **66.4** | **2** | **662** | **5.52** | **0.0042** |
| trimester * age | 23.6 | 11.8 | 2 | 619 | 0.98 | 0.3764 |
| trimester * income to needs ratio | 38.6 | 19.3 | 2 | 612 | 1.6 | 0.2022 |
| trimester * cotinine status | 73 | 36.5 | 2 | 633 | 3.03 | 0.0489 |
| CUP Status * age | 0.2 | 0.2 | 1 | 937 | 0.01 | 0.9045 |
| CUP Status * income to needs ratio | 0 | 0 | 1 | 880 | 0 | 0.9955 |
| CUP Status * cotinine status | 0.5 | 0.5 | 1 | 876 | 0.04 | 0.8438 |

**eTables 12-16: Pairwise contrasts for insomnia analyses**

| Contrast | Estimate | SE | df | t ratio | p-value |
|---|---|---|---|---|---|
| **eTable 12:** Pairwise Contrasts of Insomnia Symptom Sensitivity Analyses Adjusted for Missing T3 Insomnia Symptom Indicator | | | | | |
| **trimester1 nocan - trimester2 nocan** | **1.291** | **0.506** | **733** | **2.552** | **0.011** |
| **trimester1 nocan - trimester3 nocan** | **1.384** | **0.504** | **733** | **2.745** | **0.006** |
| trimester1 nocan - trimester1 can | 0.491 | 0.571 | 880 | 0.859 | 0.39 |
| trimester1 nocan - trimester2 can | 0.723 | 0.578 | 886 | 1.251 | 0.211 |
| trimester1 nocan - trimester3 can | -0.309 | 0.586 | 887 | -0.527 | 0.598 |
| trimester2 nocan - trimester3 nocan | 0.093 | 0.467 | 693 | 0.199 | 0.842 |
| trimester2 nocan - trimester1 can | -0.8 | 0.502 | 864 | -1.594 | 0.111 |
| trimester2 nocan - trimester2 can | -0.568 | 0.564 | 883 | -1.007 | 0.314 |
| **trimester2 nocan - trimester3 can** | **-1.6** | **0.557** | **895** | **-2.875** | **0.004** |
| trimester3 nocan - trimester1 can | -0.893 | 0.471 | 849 | -1.897 | 0.058 |
| trimester3 nocan - trimester2 can | -0.661 | 0.512 | 882 | -1.291 | 0.197 |
| **trimester3 nocan - trimester3 can** | **-1.693** | **0.553** | **887** | **-3.061** | **0.002** |
| trimester1 can - trimester2 can | 0.233 | 0.379 | 710 | 0.614 | 0.539 |
| **trimester1 can - trimester3 can** | **-0.8** | **0.399** | **718** | **-2.004** | **0.045** |
| **trimester2 can - trimester3 can** | **-1.033** | **0.416** | **688** | **-2.483** | **0.013** |
| **eTable 13:** Pairwise Contrasts of Insomnia Sensitivity Analyses Adjusted for Missing T3 CUP Status Indicator | | | | | |
| **trimester1 nocan - trimester2 nocan** | **1.29** | **0.506** | **733** | **2.55** | **0.011** |
| **trimester1 nocan - trimester3 nocan** | **1.386** | **0.503** | **736** | **2.754** | **0.006** |
| trimester1 nocan - trimester1 can | 0.496 | 0.571 | 880 | 0.869 | 0.385 |

| | | | | | |
|---|---|---|---|---|---|
| trimester1 nocan - trimester2 can | 0.73 | 0.578 | 886 | 1.263 | 0.207 |
| trimester1 nocan - trimester3 can | -0.303 | 0.586 | 890 | -0.518 | 0.605 |
| trimester2 nocan - trimester3 nocan | 0.096 | 0.466 | 695 | 0.207 | 0.836 |
| trimester2 nocan - trimester1 can | -0.794 | 0.502 | 863 | -1.581 | 0.114 |
| trimester2 nocan - trimester2 can | -0.56 | 0.564 | 883 | -0.992 | 0.321 |
| **trimester2 nocan - trimester3 can** | **-1.593** | **0.556** | **896** | **-2.863** | **0.004** |
| trimester3 nocan - trimester1 can | -0.89 | 0.47 | 855 | -1.894 | 0.059 |
| trimester3 nocan - trimester2 can | -0.656 | 0.511 | 883 | -1.283 | 0.2 |
| **trimester3 nocan - trimester3 can** | **-1.69** | **0.553** | **887** | **-3.054** | **0.002** |
| trimester1 can - trimester2 can | 0.234 | 0.379 | 711 | 0.619 | 0.536 |
| **trimester1 can - trimester3 can** | **-0.799** | **0.398** | **725** | **-2.008** | **0.045** |
| **trimester2 can - trimester3 can** | **-1.034** | **0.416** | **689** | **-2.487** | **0.013** |

**eTable 14:** Pairwise Contrasts of Insomnia Sensitivity Analyses Adjusted for Both T3 Missingness Indicators

| | | | | | |
|---|---|---|---|---|---|
| **trimester1 nocan - trimester2 nocan** | **1.289** | **0.506** | **733** | **2.548** | **0.011** |
| **trimester1 nocan - trimester3 nocan** | **1.385** | **0.504** | **732** | **2.748** | **0.006** |
| trimester1 nocan - trimester1 can | 0.495 | 0.571 | 880 | 0.867 | 0.386 |
| trimester1 nocan - trimester2 can | 0.73 | 0.578 | 885 | 1.262 | 0.207 |
| trimester1 nocan - trimester3 can | -0.304 | 0.587 | 887 | -0.519 | 0.604 |
| trimester2 nocan - trimester3 nocan | 0.096 | 0.467 | 692 | 0.206 | 0.837 |
| trimester2 nocan - trimester1 can | -0.794 | 0.502 | 863 | -1.58 | 0.114 |
| trimester2 nocan - trimester2 can | -0.56 | 0.564 | 882 | -0.992 | 0.321 |
| **trimester2 nocan - trimester3 can** | **-1.594** | **0.557** | **894** | **-2.862** | **0.004** |
| trimester3 nocan - trimester1 can | -0.89 | 0.471 | 849 | -1.889 | 0.059 |
| trimester3 nocan - trimester2 can | -0.656 | 0.512 | 881 | -1.281 | 0.2 |
| **trimester3 nocan - trimester3 can** | **-1.69** | **0.553** | **886** | **-3.054** | **0.002** |
| trimester1 can - trimester2 can | 0.234 | 0.379 | 710 | 0.618 | 0.536 |
| **trimester1 can - trimester3 can** | **-0.8** | **0.399** | **718** | **-2.003** | **0.046** |
| **trimester2 can - trimester3 can** | **-1.034** | **0.416** | **687** | **-2.486** | **0.013** |

**eTable 15:** Pairwise Contrasts of Sensitivity Analyses Restricted to Observed T3 CUP Status and Insomnia Symptoms

| | | | | | |
|---|---|---|---|---|---|
| **trimester1 nocan - trimester2 nocan** | **1.24** | **0.549** | **650** | **2.258** | **0.024** |
| **trimester1 nocan - trimester3 nocan** | **1.399** | **0.527** | **656** | **2.653** | **0.008** |
| trimester1 nocan - trimester1 can | 0.607 | 0.622 | 768 | 0.976 | 0.329 |
| trimester1 nocan - trimester2 can | 0.617 | 0.622 | 785 | 0.992 | 0.322 |
| trimester1 nocan - trimester3 can | -0.299 | 0.62 | 798 | -0.482 | 0.63 |
| trimester2 nocan - trimester3 nocan | 0.159 | 0.489 | 637 | 0.324 | 0.746 |
| trimester2 nocan - trimester1 can | -0.633 | 0.557 | 777 | -1.137 | 0.256 |
| trimester2 nocan - trimester2 can | -0.623 | 0.612 | 800 | -1.018 | 0.309 |
| **trimester2 nocan - trimester3 can** | **-1.539** | **0.593** | **821** | **-2.595** | **0.01** |
| trimester3 nocan - trimester1 can | -0.792 | 0.494 | 784 | -1.602 | 0.11 |
| trimester3 nocan - trimester2 can | -0.782 | 0.533 | 815 | -1.468 | 0.143 |
| **trimester3 nocan - trimester3 can** | **-1.698** | **0.565** | **826** | **-3.005** | **0.003** |
| trimester1 can - trimester2 can | 0.01 | 0.407 | 646 | 0.024 | 0.981 |
| **trimester1 can - trimester3 can** | **-0.906** | **0.411** | **658** | **-2.202** | **0.028** |
| **trimester2 can - trimester3 can** | **-0.916** | **0.428** | **635** | **-2.14** | **0.033** |

**eTable 16:** Pairwise Contrasts Restricted to Cannabis Use During Pregnancy Status Data Available at All Three Trimesters

| | | | | | |
|---|---|---|---|---|---|
| **trimester1 nocan - trimester2 nocan** | **1.339** | **0.537** | **651** | **2.49** | **0.013** |
| **trimester1 nocan - trimester3 nocan** | **1.734** | **0.528** | **657** | **3.28** | **0.001** |
| trimester1 nocan - trimester1 can | 0.688 | 0.611 | 766 | 1.13 | 0.261 |
| trimester1 nocan - trimester2 can | 0.859 | 0.615 | 779 | 1.4 | 0.163 |
| trimester1 nocan - trimester3 can | 0.025 | 0.62 | 786 | 0.04 | 0.968 |
| trimester2 nocan - trimester3 nocan | 0.395 | 0.485 | 638 | 0.82 | 0.415 |
| trimester2 nocan - trimester1 can | -0.651 | 0.54 | 772 | -1.21 | 0.228 |
| trimester2 nocan - trimester2 can | -0.479 | 0.598 | 793 | -0.8 | 0.423 |
| **trimester2 nocan - trimester3 can** | **-1.313** | **0.585** | **807** | **-2.25** | **0.025** |
| trimester3 nocan - trimester1 can | -1.046 | 0.492 | 774 | -2.13 | 0.034 |
| trimester3 nocan - trimester2 can | -0.875 | 0.532 | 802 | -1.64 | 0.1 |
| **trimester3 nocan - trimester3 can** | **-1.709** | **0.571** | **806** | **-2.99** | **0.003** |
| trimester1 can - trimester2 can | 0.171 | 0.395 | 645 | 0.43 | 0.664 |
| trimester1 can - trimester3 can | -0.663 | 0.411 | 657 | -1.61 | 0.108 |
| trimester2 can - trimester3 can | -0.834 | 0.427 | 634 | -1.95 | 0.051 |

**eTables 17-20 Associations with Maternal Sleep Problems and Cannabis Frequency.** False discovery rate correction was conducted for each respective variable across all four sleep phenotypes

**eTable 17:** Associations with Maternal Restless Legs Symptoms and Cannabis Frequency

| Variable | Sum Sq | Mean Sq | NumDF | DenDF | F value | p-value | p_FDR |
|---|---|---|---|---|---|---|---|
| trimester | 4.961 | 2.481 | 2 | 277.416 | 4.271 | 0.015 | - |
| cannabis frequency | 0.074 | 0.074 | 1 | 400.060 | 0.128 | 0.721 | 0.958 |
| age | 1.013 | 1.013 | 1 | 208.588 | 1.744 | 0.188 | - |
| income to need ratio | 0.235 | 0.235 | 1 | 204.982 | 0.404 | 0.526 | - |
| cotinine status | 0.008 | 0.008 | 1 | 207.954 | 0.014 | 0.905 | - |
| trimester * cannabis frequency | 0.177 | 0.089 | 2 | 333.216 | 0.152 | 0.859 | 0.859 |
| trimester * age | 0.366 | 0.183 | 2 | 273.337 | 0.315 | 0.730 | - |
| trimester * income to need ratio | 0.482 | 0.241 | 2 | 273.488 | 0.415 | 0.661 | - |
| trimester * cotinine status | 1.019 | 0.510 | 2 | 269.724 | 0.877 | 0.417 | - |
| cannabis frequency * age | 0.307 | 0.307 | 1 | 400.602 | 0.528 | 0.468 | - |
| cannabis frequency * income to need ratio | 0.025 | 0.025 | 1 | 394.475 | 0.044 | 0.835 | - |
| cannabis frequency * cotinine status | 0.335 | 0.335 | 1 | 406.718 | 0.576 | 0.448 | - |

**eTable 18:** Associations with Maternal Insomnia Symptoms and Cannabis Frequency

| Variable | Sum Sq | Mean Sq | NumDF | DenDF | F value | p-value | p_FDR |
|---|---|---|---|---|---|---|---|
| trimester | 54.67 | 27.33 | 2 | 250.53 | 2.396 | 0.093 | 0.124 |
| cannabis frequency | 7.51 | 7.51 | 1 | 360.89 | 0.658 | 0.418 | 0.835 |
| age | 14.74 | 14.74 | 1 | 198.07 | 1.292 | 0.257 | - |
| income to need ratio | 7.41 | 7.41 | 1 | 194.18 | 0.650 | 0.421 | - |
| cotinine status | 1.03 | 1.03 | 1 | 200.24 | 0.091 | 0.764 | - |

| | | | | | | | |
|---|---|---|---|---|---|---|---|
| trimester * cannabis frequency | 3.58 | 1.79 | 2 | 296.38 | 0.157 | 0.855 | 0.859 |
| trimester * age | 10.41 | 5.21 | 2 | 245.81 | 0.456 | 0.634 | - |
| trimester * income to need ratio | 44.74 | 22.37 | 2 | 244.74 | 1.961 | 0.143 | - |
| trimester * cotinine status | 32.49 | 16.24 | 2 | 243.18 | 1.424 | 0.243 | - |
| cannabis frequency * age | 0.00 | 0.00 | 1 | 361.02 | 0.000 | 0.993 | - |
| cannabis frequency * income to need ratio | 1.75 | 1.75 | 1 | 349.93 | 0.153 | 0.696 | - |
| cannabis frequency * cotinine status | 16.09 | 16.09 | 1 | 370.91 | 1.410 | 0.236 | - |

**eTable 19:** Associations with Maternal Daytime Sleepiness and Cannabis Frequency

| Variable | Sum Sq | Mean Sq | NumDF | DenDF | F value | p-value | p_FDR |
|---|---|---|---|---|---|---|---|
| trimester | 33.63 | 16.82 | 2.00 | 234.07 | 1.57 | 0.21 | - |
| cannabis frequency | 0.03 | 0.03 | 1.00 | 358.70 | 0.00 | 0.96 | 0.96 |
| age | 82.57 | 82.57 | 1.00 | 193.32 | 7.72 | 0.01 | - |
| income to need ratio | 2.93 | 2.93 | 1.00 | 183.58 | 0.27 | 0.60 | - |
| cotinine status | 70.16 | 70.16 | 1.00 | 188.97 | 6.56 | 0.01 | - |
| trimester * cannabis frequency | 3.71 | 1.85 | 2.00 | 287.32 | 0.17 | 0.84 | 0.86 |
| trimester * age | 74.94 | 37.47 | 2.00 | 237.42 | 3.50 | 0.03 | - |
| trimester * income to need ratio | 0.66 | 0.33 | 2.00 | 237.23 | 0.03 | 0.97 | - |
| trimester * cotinine status | 9.86 | 4.93 | 2.00 | 227.20 | 0.46 | 0.63 | - |
| cannabis frequency * age | 20.34 | 20.34 | 1.00 | 366.56 | 1.90 | 0.17 | - |
| cannabis frequency * income to need ratio | 28.28 | 28.28 | 1.00 | 364.81 | 2.65 | 0.10 | - |
| cannabis frequency * cotinine status | 3.60 | 3.60 | 1.00 | 361.74 | 0.34 | 0.56 | - |

**eTable 20:** Associations with Maternal Sleep Quality and Cannabis Frequency

| Variable | Sum Sq | Mean Sq | NumDF | DenDF | F value | p-value | p_FDR |
|---|---|---|---|---|---|---|---|
| trimester | 1.86 | 0.93 | 2.00 | 281.50 | 2.81 | 0.06 | - |
| cannabis frequency | 0.91 | 0.91 | 1.00 | 408.96 | 2.76 | 0.10 | 0.39 |
| age | 0.13 | 0.13 | 1.00 | 202.16 | 0.40 | 0.53 | - |
| income to need ratio | 0.26 | 0.26 | 1.00 | 194.12 | 0.77 | 0.38 | - |
| cotinine status | 0.24 | 0.24 | 1.00 | 200.78 | 0.72 | 0.40 | - |
| trimester * cannabis frequency | 0.70 | 0.35 | 2.00 | 348.54 | 1.06 | 0.35 | 0.86 |
| trimester * age | 1.23 | 0.61 | 2.00 | 280.90 | 1.85 | 0.16 | - |
| trimester * income to need ratio | 0.12 | 0.06 | 2.00 | 276.53 | 0.18 | 0.83 | - |

| | | | | | | | |
|---|---|---|---|---|---|---|---|
| trimester * cotinine status | 0.11 | 0.06 | 2.00 | 275.92 | 0.17 | 0.84 | - |
| cannabis frequency * age | 0.95 | 0.95 | 1.00 | 411.52 | 2.86 | 0.09 | - |
| cannabis frequency * income to need ratio | 0.24 | 0.24 | 1.00 | 401.85 | 0.72 | 0.40 | - |
| cannabis frequency * cotinine status | 0.05 | 0.05 | 1.00 | 413.60 | 0.14 | 0.71 | - |

**eTables: 21-24 Associations With Maternal Sleep Problems and Motivation for Use.**

**eTable 21:** Associations With Maternal Restless Legs Symptoms and Motivations for Use

| **Variable** | **Sum Sq** | **Mean Sq** | **NumDF** | **DenDF** | **F value** | **p-value** | **p_FDR** |
|---|---|---|---|---|---|---|---|
| trimester | 6.15 | 3.074 | 2 | 353 | 5.42 | 0.005 | - |
| motivation for use | 1.45 | 1.452 | 1 | 241 | 2.56 | 0.111 | 0.148 |
| age | 1.55 | 1.548 | 1 | 249 | 2.73 | 0.100 | - |
| income to needs ratio | 0.71 | 0.714 | 1 | 256 | 1.26 | 0.263 | - |
| cotinine status | 0.08 | 0.079 | 1 | 243 | 0.14 | 0.709 | - |
| trimester * motivation for use | 0.22 | 0.108 | 2 | 337 | 0.19 | 0.827 | 0.827 |
| trimester * age | 1.01 | 0.503 | 2 | 349 | 0.89 | 0.413 | - |
| trimester * income to needs ratio | 0.64 | 0.322 | 2 | 366 | 0.57 | 0.568 | - |
| trimester * cotinine status | 2.66 | 1.328 | 2 | 343 | 2.34 | 0.098 | - |
| motivation for use* age | 0.38 | 0.379 | 1 | 242 | 0.67 | 0.414 | - |
| motivation for use* income to needs ratio | 0.02 | 0.022 | 1 | 252 | 0.04 | 0.843 | - |
| motivation for use* cotinine status | 0.88 | 0.877 | 1 | 239 | 1.55 | 0.215 | - |

**eTable 22:** Associations With Insomnia Symptoms and Motivations for Use

| **Variable** | **Sum Sq** | **Mean Sq** | **NumDF** | **DenDF** | **F value** | **p-value** | **p_FDR** |
|---|---|---|---|---|---|---|---|
| trimester | 45.1 | 22.5 | 2 | 297 | 2.11 | 0.123 | - |
| **motivation for use** | **139.7** | **139.7** | **1** | **223** | **13.09** | **3.70E-04** | **0.001** |
| age | 8.1 | 8.1 | 1 | 228 | 0.76 | 0.385 | - |
| income to needs ratio | 9.4 | 9.4 | 1 | 226 | 0.88 | 0.348 | - |
| cotinine status | 3.9 | 3.9 | 1 | 228 | 0.37 | 0.546 | - |
| trimester * motivation for use | 34.6 | 17.3 | 2 | 292 | 1.62 | 0.200 | 0.399 |
| trimester * age | 2 | 1 | 2 | 300 | 0.09 | 0.911 | - |
| trimester * income to needs ratio | 54 | 27 | 2 | 298 | 2.53 | 0.081 | - |
| trimester * cotinine status | 31.9 | 16 | 2 | 296 | 1.5 | 0.226 | - |
| motivation for use* age | 12.5 | 12.5 | 1 | 224 | 1.17 | 0.280 | - |
| motivation for use* income to needs ratio | 3.4 | 3.4 | 1 | 228 | 0.32 | 0.574 | - |

| | | | | | | | |
|---|---|---|---|---|---|---|---|
| motivation for use* cotinine status | 21.9 | 21.9 | 1 | 225 | 2.05 | 0.153 | - |

**eTable 23:** Associations With Maternal Daytime Sleepiness and Motivation for Use

| Variable | Sum Sq | Mean Sq | NumDF | DenDF | F value | p-value | p_FDR |
|---|---|---|---|---|---|---|---|
| trimester | 19.2 | 9.6 | 2 | 260 | 0.95 | 0.39 | - |
| motivation for use | 19.2 | 19.2 | 1 | 204 | 1.89 | 0.171 | 0.171 |
| age | 36.5 | 36.5 | 1 | 214 | 3.59 | 0.059 | - |
| income to needs ratio | 0 | 0 | 1 | 206 | 0 | 0.965 | - |
| cotinine status | 50.1 | 50.1 | 1 | 209 | 4.93 | 0.027 | - |
| trimester * motivation for use | 64.1 | 32 | 2 | 256 | 3.15 | 0.044 | 0.178 |
| trimester * age | 55.3 | 27.7 | 2 | 269 | 2.72 | 0.068 | - |
| trimester * income to needs ratio | 14.1 | 7.1 | 2 | 264 | 0.69 | 0.5 | - |
| trimester * cotinine status | 14.1 | 7.1 | 2 | 262 | 0.7 | 0.5 | - |
| motivation for use* age | 2.6 | 2.6 | 1 | 210 | 0.25 | 0.616 | - |
| motivation for use* income to needs ratio | 5.1 | 5.1 | 1 | 210 | 0.5 | 0.478 | - |
| motivation for use* cotinine status | 0.4 | 0.4 | 1 | 206 | 0.04 | 0.85 | - |

**eTable 24:** Associations With Maternal Sleep Quality and Motivation for Use

| Variable | Sum Sq | Mean Sq | NumDF | DenDF | F value | p-value | p_FDR |
|---|---|---|---|---|---|---|---|
| trimester | 2.295 | 1.148 | 2 | 322 | 3.75 | 0.025 | - |
| **motivation for use** | **2.407** | **2.407** | **1** | **219** | **7.86** | **0.006** | **0.011** |
| age | 0.06 | 0.06 | 1 | 226 | 0.2 | 0.659 | - |
| income to needs ratio | 0.369 | 0.369 | 1 | 225 | 1.2 | 0.274 | - |
| cotinine status | 0.263 | 0.263 | 1 | 223 | 0.86 | 0.355 | - |
| trimester * motivation for use | 0.297 | 0.148 | 2 | 315 | 0.48 | 0.616 | - |
| trimester * age | 1.229 | 0.615 | 2 | 326 | 2.01 | 0.136 | - |
| trimester * income to needs ratio | 0.211 | 0.106 | 2 | 326 | 0.34 | 0.709 | - |
| trimester * cotinine status | 0.004 | 0.002 | 2 | 320 | 0.01 | 0.993 | - |
| motivation for use* age | 0.202 | 0.202 | 1 | 219 | 0.66 | 0.418 | - |
| motivation for use* income to needs ratio | 1.454 | 1.454 | 1 | 227 | 4.75 | 0.030 | - |
| motivation for use* cotinine status | 0.248 | 0.248 | 1 | 220 | 0.81 | 0.369 | - |

**eTables 25-26:** Motivation for Use Pairwise Contrasts.

**eTable 25:** Pairwise Contrasts for Motivation for Use and Insomnia Symptoms

| Result type | Motivation for use /Contrast | Emmean/Estimate | SE | df | Lower CL | Upper CL | t ratio | p-value |
|---|---|---|---|---|---|---|---|---|

| | | | | | | | | |
|---|---|---|---|---|---|---|---|---|
| Estimated mean | other | 7.35 | 0.41 | 238 | 6.55 | 8.16 | - | - |
| Estimated mean | sleep | 9.53 | 0.505 | 222 | 8.53 | 10.52 | - | - |
| **Contrast** | **other - sleep** | **-2.18** | **0.65** | **228** | **-** | **-** | **-3.35** | **9.00E-04** |

**eTable 26:** Pairwise Contrasts for Motivation for Use and Sleep Quality

| Result type | Motivation for use/Contrast | Emmean/Estimate | SE | df | Lower CL | Upper CL | t ratio | p-value |
|---|---|---|---|---|---|---|---|---|
| Estimated mean | other | 2.14 | 0.0521 | 236 | 2.03 | 2.24 | - | - |
| Estimated mean | sleep | 2.29 | 0.0633 | 212 | 2.16 | 2.41 | - | - |
| Contrast | other - sleep | -0.154 | 0.0819 | 221 | - | - | -1.875 | 0.0621 |
